# Mapping the landscape: A protocol of a jurisdictional scan of self-identified learning health systems

**DOI:** 10.1101/2023.10.26.23297605

**Authors:** Shelley Vanderhout, Marissa Bird, Antonia Giannarakos, Carly Whitmore

## Abstract

**Background:** There is a growing movement to implement learning health systems (LHS), in which real-time evidence, informatics, patient-provider partnerships and experiences, and organizational culture are aligned to support improvements in care. However, what constitutes a LHS varies based on context and capacity, hindering standardization, scale-up, and knowledge sharing. Further, LHS often use “usual care” as the benchmark for comparing new approaches to care, but disentangling usual care from multifarious care modalities found across settings is challenging. To advance robust LHS, a comprehensive overview of existing LHS including strengths and opportunities for growth is needed.

**Objectives:** To scope and identify international existing LHS to: 1) inform the global landscape of LHS, highlight common strengths, and identify opportunities for growth or improvement; and 2) identify common characteristics, emphases, assumptions, or challenges described in establishing counterfactuals in LHS.

**Methods:** A jurisdictional scan will be conducted according to modified PRISMA guidelines. LHS will be identified through a search of peer-reviewed and grey literature using Ovid Medline, Ebsco CINAHL, Ovid Embase, Clarivate Web of Science, and PubMed Non-Medline databases and the web along with informal discussions with peer LHS experts. Self-identified LHS will be included if they are described in sufficient detail, either in literature or during informal discussions, according to ≥4 of 10 criteria (core functionalities, analytics, use of evidence, co-design/implementation, evaluation, change management/governance structures, data sharing, knowledge sharing, training/capacity building, equity, sustainability) in an existing framework to characterize LHS. Search results will be screened, extracted, and analyzed to inform two descriptive reviews pertaining to our two main objectives. Data will be extracted according to a pre-specified extraction form and summarized descriptively.

**Implications:** This research will characterize the current landscape of worldwide LHS and provide a foundation for promoting knowledge and resource sharing, identifying next steps for the growth, improvement, and evaluation of LHS.

## Introduction

Evidence-to-practice gaps in health care contribute to inefficient and ineffective care, ballooning costs, poor experiences for patients, frustration and burnout for healthcare providers, and widening health inequities.^1^ In response to these challenges, there is a growing emphasis on the development, implementation, and evaluation of innovative approaches, such as learning health systems (LHS). LHS have been developed to address gaps between knowledge and practice by combining real-time evidence, informatics systems, patient-provider partnerships and experiences, and institutional strategies to support continuous innovation and improvements in care.^2^ Worldwide, a handful of groups and institutions have recognized the potential for LHS to transform the status quo, but these tend to be specialized in focus, siloed, and inconsistently consider equity in their approach.^3^ Furthermore, existing LHS are incomprehensively characterized in the literature, leading to gaps in knowledge transfer, resource sharing, and the development of best practices.

LHS often set out to embed cycles of data collection, knowledge synthesis, and practice change,^4^ using “usual care” as the benchmark or counterfactual for comparing new interventions or approaches to care. However, in the evolving landscape of pragmatic and realist research, teasing apart what is ‘usual’ has become a challenging endeavour. This is particularly a challenge when attempting to disentangle new care solutions from the multifarious existing care modalities found across settings. As a result, empirical evidence has occasionally revealed unexpected outcomes when comparing complex care models or approaches to usual care, wherein, for example, the introduction of novel elements, such as healthcare worker interventions featuring more frequent touchpoints and early health challenge detection, can paradoxically result in adverse outcomes, such as increased hospital admissions.^5,6^ Assessing the impact of new care models in a LHS should be assessed holistically, including investigation into not just ‘what works’, but inclusions of questions about, ‘for whom, under which circumstances, and why?’.

Despite best efforts to share methodological innovations and lessons learned specific to LHS, a comprehensive scan of existing LHS worldwide and thorough description of their characteristics including evaluation approaches, is needed. Characterizing LHS and relevant contextual factors will provide a foundation for developing methodological advances, sharing knowledge, and unraveling the intricate web of factors influencing the effectiveness and impact of innovative care systems.

In this protocol, we describe a jurisdictional scan to scope and characterize international examples of self-identified LHS. The resulting sample of LHS we identify will then be used to conduct two descriptive reviews with the following objectives, respectively:

1. To scope and identify international examples of self-identified LHS and characterize the identified systems according to an existing framework.
2. To elucidate common characteristics, emphases, assumptions, or challenges described in establishing counterfactuals in learning health system research, and describe how these counterfactuals impact the evaluation process with a focus on health equity.

Our research approach will use the same search strategy to identify studies for these two reviews. The focus of this jurisdictional scan will be intentionally broad, aimed at capturing the most salient issues in LHS in practice. In the nested counterfactuals review, we will specifically attend to measurement and evaluation issues in Learning Health Systems, a topic that has remained an area of considerable debate.^7^

## Materials and methods

Jurisdictional scans are used to explore and understand how problems have been framed by others in a given field, and to compare and assess the strengths and weaknesses of each approach.^8^ In addition, they aim to produce policy-relevant results by including detail of the problem in context, making them useful tools for understanding how a specific initiative has been framed, conducted, or disseminated in other jurisdictions.^9^ From these insights, key learnings, challenges, and implementation considerations may be distilled. This jurisdictional scan will use two approaches to identify potentially relevant LHS: a literature review and informal discussions. The resulting sample of eligible LHS will then be included in two descriptive reviews corresponding to Objectives 1 and 2. This study has been registered on the Open Science Framework at osf.io/b5u7e.

### Theoretical Approach

The LHS Action Framework^10^ (Figure 1) will be used to map LHS characteristics. This framework was developed by consolidating existing LHS frameworks in the literature, and describes how research and health care operations are linked and enacted in a comprehensive LHS approach to advance population health and health equity. It was produced to identify capabilities necessary to enact the learning elements required, including key questions and methods, to ensure a systematic approach to learning and achieve equity-centered quadruple aim metrics.

**Figure 1.**
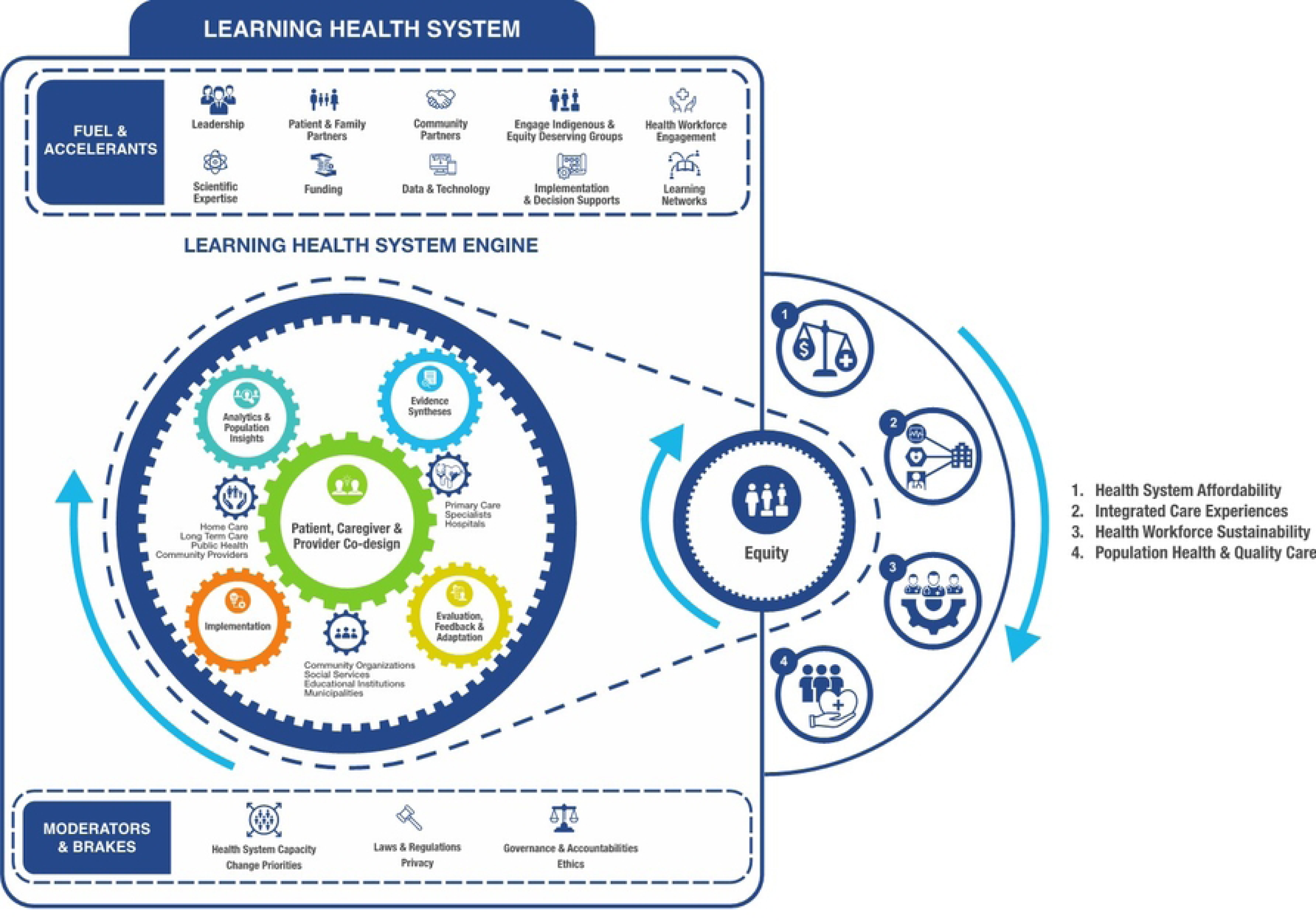
Learning Health System Action Framework.

The LHS Action Framework has five learning gears: analytics and population insights, evidence syntheses, patient, caregiver and provider co-design, implementation, and evaluation, and three health system gears representing different care settings, services, and institutions. Central to this framework is the outcome of improved equity, such that all LHS activities are built on principles of inclusivity, accessibility, the value created for equity-deserving groups.

This model will also be used to characterize LHS counterfactuals, including the extent of LHS implementation, adaptations made to a LHS as it matures, unintended consequences, and process indicators of success.^11–13^

### Literature Review

In absence of specific guidance for jurisdictional scans, we adapted the Preferred Reporting Items for Systematic Reviews and Meta-Analyses Extension for Scoping Reviews (PRISMA-ScR) checklist (see Supporting Information: PRISMA-ScR Checklist). An information specialist (AG) supported the search development and implementation across Ovid Medline, Ebsco CINAHL, Ovid Embase, Clarivate Web of Science, and PubMed Non-Medline databases, as well as grey literature and relevant websites (see Supporting Information: Search strategy). We built upon the search strategy used by Enticott et al.^3^ and define LHS according to the Institute of Medicine’s^14^ definition: “a system in which progress in science, informatics, and care culture align to generate new knowledge as an ongoing, natural by-product of the care experience, and seamlessly refine and deliver best practices for continuous improvement in health and healthcare.” We will also leverage professional networks to identify relevant sources of literature in the form of websites, newsletters, or online or print reports.

### Informal discussions

We will conduct informal virtual discussions to complement the literature search in two ways: first, to facilitate data extraction for new LHS not identified in the literature search, and second, to complement existing information from the literature search where appropriate. We will consult our professional networks to identify individuals who are key stakeholders in existing LHS and may be able to provide us relevant information, and identify corresponding authors of publications identified in the literature search to produce a list of potential participants. We will invite these individuals via email (see Supporting Information: Discussion recruitment email) to participate in these discussions (see Supporting Information: Discussion Guide). Discussions will be 30-60 minutes in length and conducted over Zoom by authors SV, CW, or MB. They will not be recorded and no personal or demographic information will be documented, but detailed notes will be taken to facilitate data extraction.

### Eligibility criteria

We will include LHS that are self-described as such in peer reviewed journal articles, reports, web pages, and informal discussions (see above). Authors or discussion participants must self-identify their LHS using at least one of the following descriptors:^13^

– Learning health[care] system

– Learning health[care] network

– Learning collaborative

– Learning laboratory

– Community-clinician participatory data healthcare research

– Data driven improvement initiative

– Practice-based data/research network

– Circular data-driven healthcare

– Rapid learning health system

Ideally, literature will be descriptive in nature, but articles describing empirical research of any design or objective in the context of a LHS will be considered. We will only include literature published after 2007, when LHS were formally conceptualized,^15^ and those written in English. Regardless of whether a LHS is identified through the literature search or discussions, it will only be included when described in sufficient detail according to at least 4 of the 10 criteria described below (see Data Extraction) to facilitate a fulsome description of the system within the resultant papers. Therefore, a LHS described only as a research setting, for example, but without further information will not be included.

Three independent screeners will review all titles and abstracts identified in the literature search in Covidence in duplicate. Authors SV, CW, and MB will resolve any uncertainties and verify the final sample.

### Data extraction

For Objective 1, each identified LHS will be described according to the following core characteristics, where data are available, from the literature search and/or informal discussions:

– LHS core functionalities

– Analytic strategies

– Use of evidence

– Co-design and implementation approaches

– Evaluation and research integration

– Change management and governance structures

– Data and infrastructure sharing processes

– Knowledge sharing practices

– Training and capacity building approaches

– Health equity considerations

– Sustainability

Additional descriptors, where available, will include:

– Country

– Public/private funding

– EMR details (Epic/other, patient portal, research integration, etc.)

– Patient population(s) served

– Patient population size

– Years since implementation

– Personnel involved (e.g., patients, health care providers, operational staff, leadership)

– Model aims

– Reference to grounding framework or model

### Analysis

Data analysis for this review will follow a two-stage process aimed at addressing our two research objectives: 1) to characterize international examples of LHS according to the LHS Action Framework,^10^ and 2) to determine common facilitators and challenges described in establishing counterfactuals in LHS research. For Objective 1, we will use a combination of quantitative (i.e., frequency counts) and qualitative (i.e., thematic analysis) methods to accurately describe the points of emphasis in global LHS and their evaluation approaches. Data from extraction tables will be merged with informal discussion data to describe models in a fulsome way. We anticipate that this analysis process may also yield gaps in the literature where future work can be focused in order to strengthen the field of LHS. For Objective 2, analysis will focus on the “Evaluation, Feedback, and Adaptation” wheel of the LHS Action Framework.^10^

See Supporting Information for the data extraction form. We will extract all data into a spreadsheet via Airtable^16^ and summarize descriptors as appropriate.

This study has been reviewed by the Research Ethics Board at Trillium Health Partners and was provided an exemption from approval, as it was deemed to be quality improvement.

### Patient Partnership and Knowledge Translation Plan

Using Graham’s Knowledge-to-Action framework,^17^ we will develop an active knowledge translation plan by: 1) identifying key messages arising from this scan; 2) determining the principal target audience for each of these messages; 3) seeking and involving the most credible messenger for these messages, and; 4) launching a knowledge translation strategy that is grounded in the best available evidence. Drawing upon a diverse range of approaches to disseminate the results of this scan, including a virtual symposium that will bring together the key target audience of this research, these strategies will ensure that these findings reflect the needs of the end-users of this information, and facilitate appropriate sharing of outputs.

### Anticipated Challenges

We foresee some potential challenges related to this jurisdictional scan. First, the yield of the literature search may be more extensive than anticipated. To overcome this, we will work closely with the information specialist to ensure that the scope of the scan is manageable but comprehensive. Second, it is anticipated that for many LHS, it will be challenging to discern or categorize characteristics solely from what is described and published. For this reason, and drawing upon the expertise and networks of our team, we will hold informal discussions with LHS leaders to comprehensively, but not systematically, scan this literature.

## Discussion

This jurisdictional scan will serve to map and characterize existing self-identified international LHS and provide an improved understanding of how these LHS describe, if at all, their counterfactuals. Knowledge generated from this research will create a needed foundation for creating a harmonized network of LHS leaders and collaborators, and highlight common characteristics of existing LHS and opportunities for growth. This work will build on an existing comprehensive framework^10^ to describe LHS, which will help inform the criteria health system leaders use to benchmark progress of a maturing LHS and set context-relevant targets for growth and improvement. Using this framework as a common language may also encourage consistent reporting, facilitate knowledge and resource sharing, streamline collaboration, and lead to refinement and new iterations of the framework to provide guidance to individuals, teams, and systems who are adopting an LHS approach.

## Author contributions

Shelley Vanderhout, Marissa Bird, and Carly Whitmore conceptualized the study, drafted the manuscript, and approved the final manuscript as submitted. Antonia Giannarakos created and implemented the search strategy.

## Data Availability

No datasets were generated or analysed during the current study. All relevant data from this study will be made available upon study completion.

## Supporting Information Captions

**S1 PRISMA-ScR Checklist**

**S2 Search strategy**

**S3 Discussion recruitment email S4 Discussion guide**

**S5 Data extraction form**

